# The fading of the mpox outbreak among men who have sex with men: a mathematical modelling study

**DOI:** 10.1101/2023.01.31.23285294

**Authors:** Maria Xiridou, Fuminari Miura, Philippe Adam, Eline Op de Coul, John de Wit, Jacco Wallinga

**Affiliations:** Centre for Infectious Disease Control, National Institute of Public Health and the Environment, Bilthoven, the Netherlands; Center for Marine Environmental Studies, Ehime University, Ehime, Japan; Centre for Social Research in Health, UNSW Sydney, Sydney, NSW, Australia; Institute for Prevention and Social Research, Utrecht, the Netherlands; Interdisciplinary Social Science, Utrecht University, Utrecht, the Netherlands; Department of Biomedical Data Sciences, Leiden University Medical Center (LUMC), Leiden, the Netherlands

**Keywords:** monkeypox, mpox, men who have sex with men, MSM, mathematical model, immunity, vaccination, sexual behaviour

## Abstract

**Background:** In the Netherlands, the number of mpox cases started declining before mpox vaccination was initiated. Most cases were men who have sex with men (MSM). We investigated whether the decline in mpox could be attributed to infection-induced immunity or behavioural adaptations.

**Methods:** We developed a transmission model and accounted for possible behavioural adaptations: less casual partners and shorter time until MSM with mpox refrain from sexual contacts.

**Results:** Without behavioural adaptations, the peak in modelled cases matched observations, but the decline was less steep than observed. With behavioural adaptations in the model, we found a decline of 16-18% in numbers of casual partners in June and 13-22% in July 2022. Model results showed a halving of the time before refraining from sex. When mpox vaccination started, 57% of MSM with very high sexual activity in the model had been infected. Model scenarios revealed that the outbreak could have waned by November 2022 even without vaccination.

**Conclusions:** The limited duration of the mpox outbreak in the Netherlands can be ascribed primarily to infection-induced immunity among MSM with high sexual activity levels. The decline was accelerated by behavioural adaptations. Immunity among those most sexually active is essential to impede mpox resurgence.

**Short summary:** Modelling study shows that the decline in mpox cases among MSM in the Netherlands was primarily due to infection-induced immunity among those with high sexual activity levels and accelerated by behavioural adaptions. The outbreak could have faded even without vaccination.

## Introduction

Mpox (formerly known as monkeypox) is a zoonotic disease caused by the monkeypox virus (MPXV) [1]. In May-July 2022, mpox outbreaks were reported, mostly among men who have sex with men (MSM) [2, 3]. The number of cases peaked within 2-3 months in most countries and then declined [2]. Several factors may have contributed to the abrupt decline, including increased immunity due to MPXV infection, behavioural adaptations, and public health interventions [3-5].

In the Netherlands, the number of mpox cases increased sharply in June 2022, but started declining in the second week of July 2022 [6], about two weeks before the national mpox vaccination programme started on 25 July 2022 [7]. In May and June 2022, a large media coverage of the mpox outbreak contributed to raise awareness of a new threat among MSM. In June 2022, health communication messages about mpox were disseminated in the media [7, 8]. Also, it was recommended to refrain from sexual or physical contacts when mpox was suspected or diagnosed [9]. In an online survey carried out in August 2022, most participants reported to be willing to refrain from close physical and sexual contacts in case of MPXV infection and some had reduced their number of sexual partners since the start of the outbreak [9]. Sex venues and parties in Amsterdam experienced a considerable drop in numbers of visitors in July 2022 [10].

Our study aimed to assess the factors that may have led to the decline in the number of mpox cases observed in the Netherlands before the start of the mpox vaccination programme. We developed a mathematical model that describes the transmission of MPXV via sexual or intimate contacts among MSM and fitted the model to numbers of mpox cases in the Netherlands until 25 July 2022, when the mpox vaccination programme started. To disentangle the impact of immunity due to infection and the impact of behavioural adaptations from the impact of vaccination in shaping the course of the outbreak, we examined also model scenarios without the vaccination programme until the end of 2022.

## Methods

### The transmission model

We developed a deterministic compartmental model that describes MPXV transmission among MSM (Figure 1). The model parameters are defined in Tables 1-3. We accounted only for transmission via sexual or intimate contacts with either main regular sex partners or casual sex partners, a distinction that follows earlier modelling of sexual relationships [11-13]. In this paper, the notion of “main regular sex partner” refers to a person with whom an individual engages in regular or ongoing sexual activity, typically (but not exclusively) within a committed relationship. Other relationships involving sexual contacts are referred to as casual partnerships in this study. Considering sexual or intimate contacts sufficient for MPXV transmission, for simplicity, we assumed one such contact per casual partner. In the model, MSM were divided into four groups, based on level of sexual activity, that was measured in terms of the total number of sex partners men have: very low, fairly low, fairly high, and very high sexual activity (Table 2).

**Table 1.**
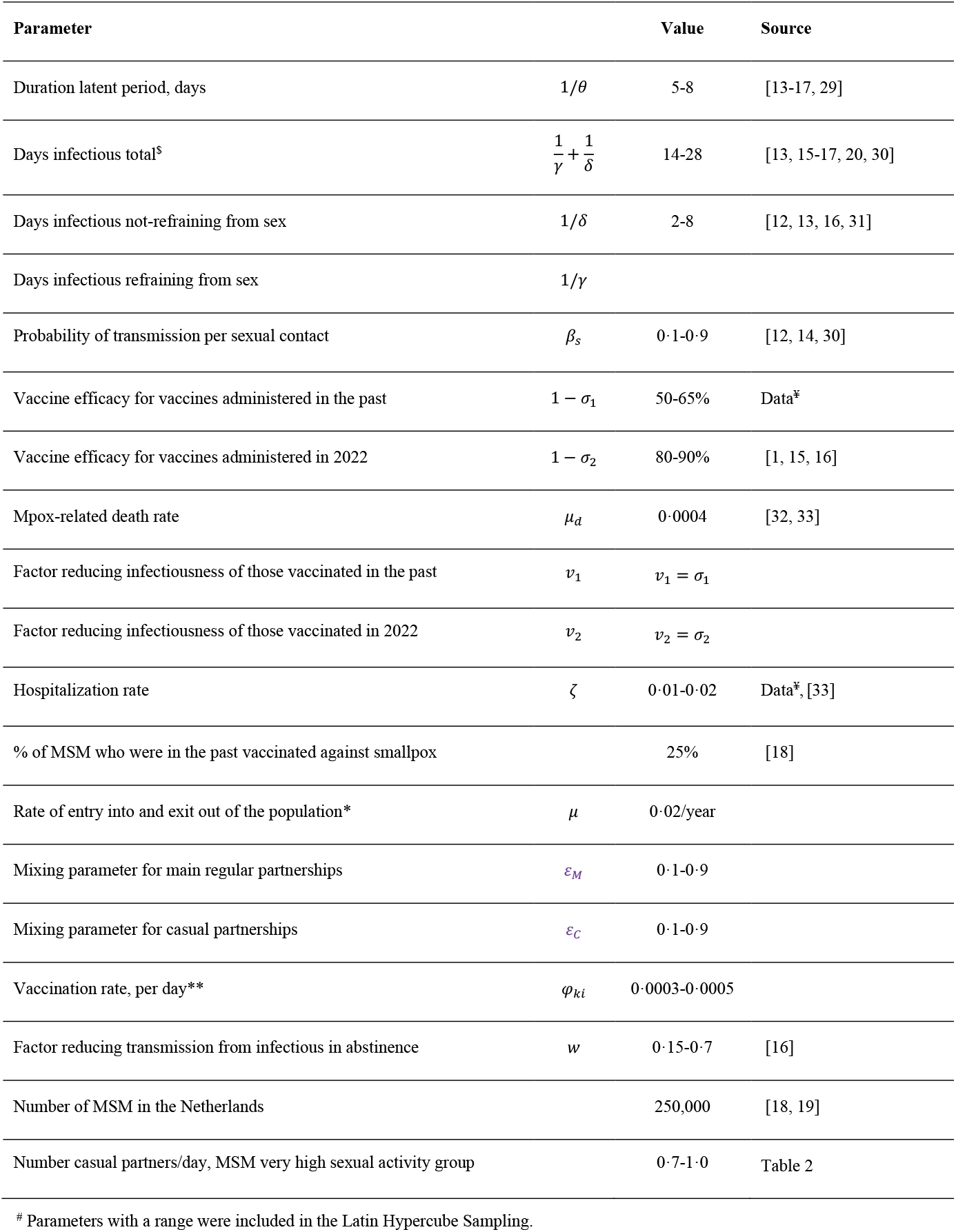
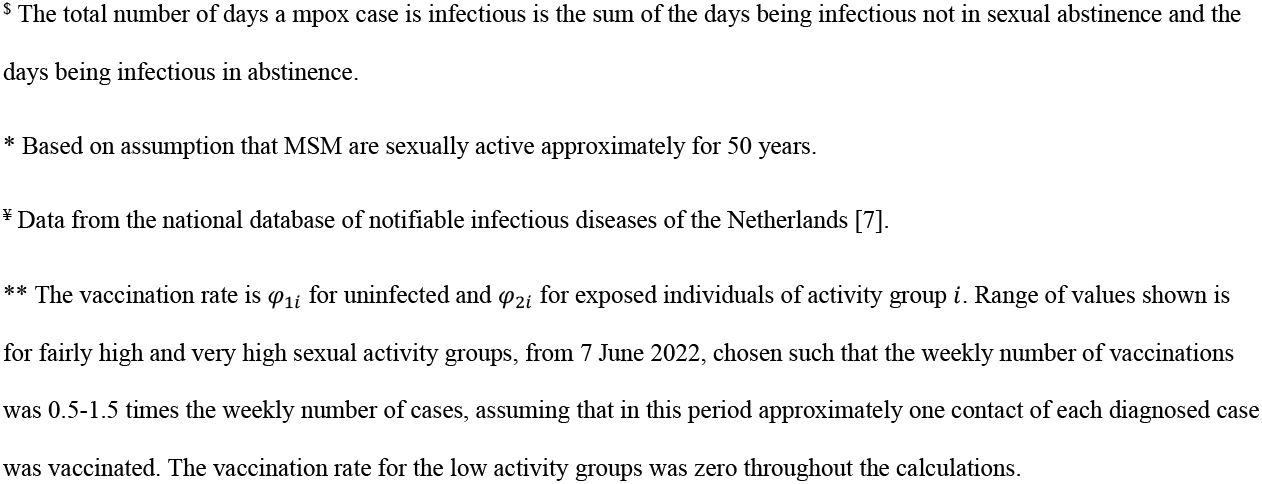
Model parameters^#^.

| Parameter |  | Value | Source |
| --- | --- | --- | --- |
| Duration latent period, days | $1/\theta$ | 5-8 | [13-17, 29] |
| Days infectious total<sup>s</sup> | $\frac{1}{\gamma} + \frac{1}{\delta}$ | 14-28 | [13, 15-17, 20, 30] |
| Days infectious not-refraining from sex | $1/\delta$ | 2-8 | [12, 13, 16, 31] |
| Days infectious refraining from sex | $1/\gamma$ | | |
| Probability of transmission per sexual contact | $\beta_s$ | 0.1-0.9 | [12, 14, 30] |
| Vaccine efficacy for vaccines administered in the past | $1 - \sigma_1$ | 50-65% | Data<sup>‡</sup> |
| Vaccine efficacy for vaccines administered in 2022 | $1 - \sigma_2$ | 80-90% | [1, 15, 16] |
| Mpox-related death rate | $\mu_d$ | 0.0004 | [32, 33] |
| Factor reducing infectiousness of those vaccinated in the past | $v_1$ | $v_1 = \sigma_1$ | |
| Factor reducing infectiousness of those vaccinated in 2022 | $v_2$ | $v_2 = \sigma_2$ | |
| Hospitalization rate | $\zeta$ | 0.01-0.02 | Data<sup>‡</sup>, [33] |
| % of MSM who were in the past vaccinated against smallpox |  | 25% | [18] |
| Rate of entry into and exit out of the population* | $\mu$ | 0.02/year | |
| Mixing parameter for main regular partnerships | $\varepsilon_M$ | 0.1-0.9 | |
| Mixing parameter for casual partnerships | $\varepsilon_C$ | 0.1-0.9 | |
| Vaccination rate, per day** | $\varphi_{ki}$ | 0.0003-0.0005 | |
| Factor reducing transmission from infectious in abstinence | $w$ | 0.15-0.7 | [16] |
| Number of MSM in the Netherlands |  | 250,000 | [18, 19] |
| Number casual partners/day, MSM very high sexual activity group |  | 0.7-1.0 | Table 2 |

<sup>#</sup> Parameters with a range were included in the Latin Hypercube Sampling.
<sup>§</sup> The total number of days a mpox case is infectious is the sum of the days being infectious not in sexual abstinence and the days being infectious in abstinence.
\* Based on assumption that MSM are sexually active approximately for 50 years.
<sup>¥</sup> Data from the national database of notifiable infectious diseases of the Netherlands [7].
\*\* The vaccination rate is $\varphi_{1i}$ for uninfected and $\varphi_{2i}$ for exposed individuals of activity group $i$ . Range of values shown is for fairly high and very high sexual activity groups, from 7 June 2022, chosen such that the weekly number of vaccinations was 0.5-1.5 times the weekly number of cases, assuming that in this period approximately one contact of each diagnosed case was vaccinated. The vaccination rate for the low activity groups was zero throughout the calculations.

**Table 2.** Parameters that depend on the sexual activity group of individuals.

| Parameter for activity group $i$ | Activity group | | | |
| --- | --- | --- | --- | --- |
|  | Very | Fairly | Fairly | Very |
|  | low | low | high | high |
| % in specific activity group* | 45.1% | 36.4% | 17.7% | 0.7% |
| Rate of forming main regular partnerships per year* ($\alpha_{Mi}$) | 0.1 | 0.2 | 0.7 | 0.7 |
| Number of sex contacts per main regular partner per day ($u_i$) | 0.03 | 0.06 | 0.14 | 0.14 |
| % with main regular partner* ($q_i$) | 60% | 51% | 54% | 42% |
| Number of casual partners per day* ($\alpha_{ci}$) | 0.006 | 0.044 | 0.139 | 0.7-1.0 |

\* Parameters estimated from data from the first round of the “COVID-19, Sex, and Intimacy Survey” [24]. We used only data from the questions relating to sexual activity in the second half of 2019 (before the start of the pandemic in the Netherlands). Men in the very high sexual activity group reported more than 100 partners in the second half of 2019.

**Figure 1.**
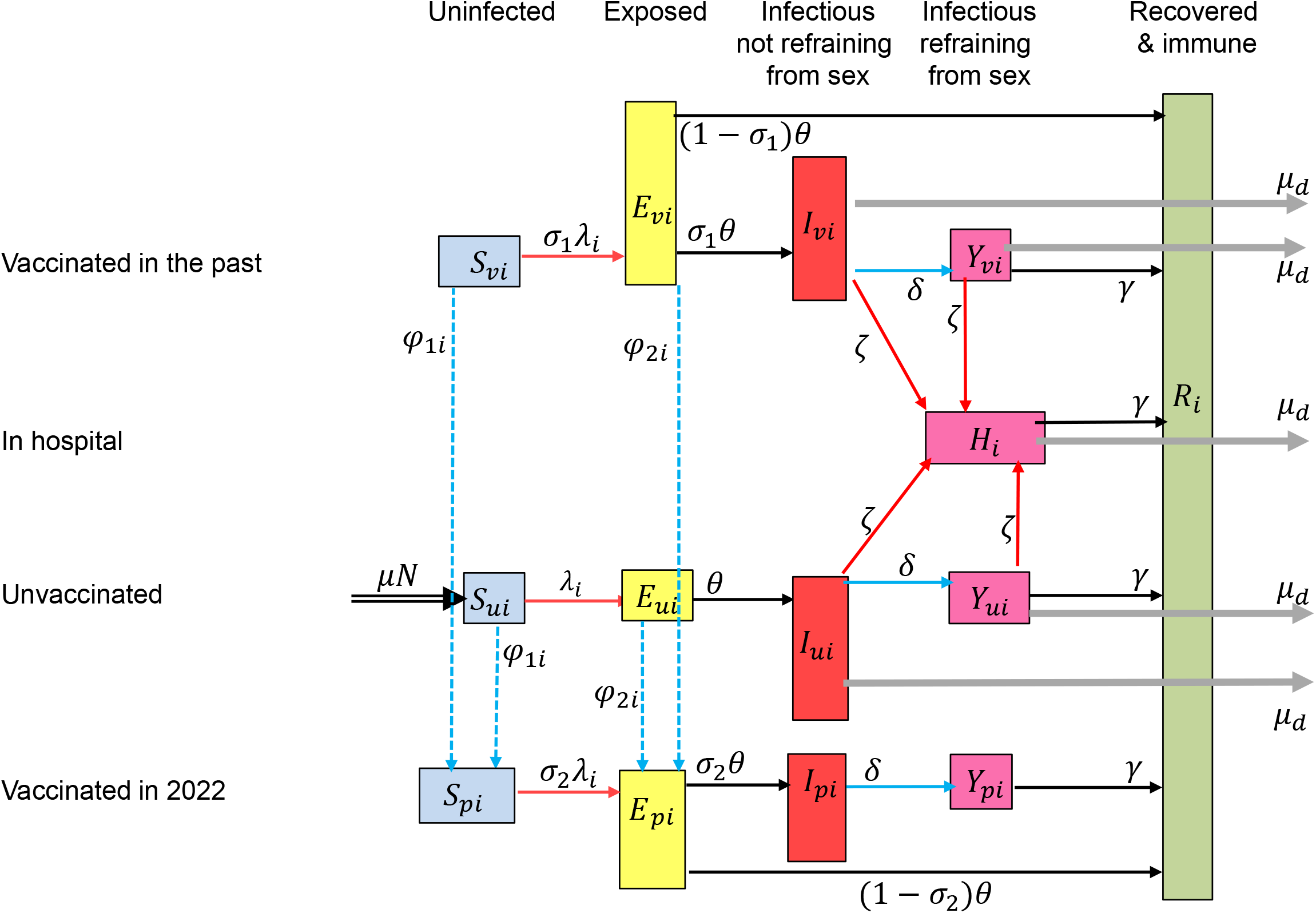
Flow diagram of the model for monkeypox virus (MPXV) infections. Individuals in the model are divided into classes according to stage of mpox infection, vaccination status, and whether they are refraining from sexual contacts. Unvaccinated individuals can be susceptible to MPXV (*S*_*ui*_), exposed to MPXV (*E*_*ui*_), infectious not refraining from sexual contacts (*I*_*ui*_), or infectious but refraining from sexual contacts (*Y*_*ui*_). Individuals vaccinated in the past can be susceptible (*S*_*vi*_), exposed (*E*_*vi*_), infectious not refraining or refraining from sexual contacts (*I*_*vi*_, *Y*_*vi*_). Individuals vaccinated in 2022 can be susceptible (*S*_*pi*_), exposed (*E*_*pi*_), infectious not refraining or refraining from sexual contacts (*I*_*pi*_, *Y*_*pi*_). Infectious cases may need hospitalization (*H*_*i*_). After recovery, they are immune (*R*_*i*_). These classes are further divided into four subclasses according to level of sexual activity. In the model, we accounted for exit out of the population at a per capita rate *μ* that is not shown in the diagram.

### The course of mpox infection

After infection, individuals are initially exposed but not yet infectious; later, they become infectious, but may not (yet) have symptoms [14, 15]. In the model, we assumed that infectious individuals may start refraining from sexual or intimate contacts a few days after becoming infectious, when they have symptoms or test positive for MPXV [15, 16]. MSM refraining from sexual/intimate contacts can still transmit MPXV (due to imperfect adherence or because they refrain only from specific types of contacts), but with a lower probability, compared to those not refraining from sexual/intimate contacts [16]. Most mpox cases recover after a few weeks and become immune [17].

### Vaccinated MSM

Smallpox vaccination was ended in the Netherlands in 1975. Based on the fraction of the male population 17-66 years old in the Netherlands being born before 1975, in the model we assumed that 25% of MSM in 2022 had been vaccinated in the past and that still offers some protection against mpox [7]. As we had no data to establish the percentage vaccinated in the past in each sexual activity group separately, we assumed the same percentage (25%) in all activity groups. The importance of this assumption was examined in sensitivity analyses (see paragraph below). From the second week of June 2022, close contacts of individuals diagnosed with mpox were offered vaccination with a single dose of Imvanex®. To account for these vaccinations, we included in the model a small vaccination rate from 8 June 2022 onwards among those with the highest level of sexual activity.

### Data

We used data on the daily numbers of mpox cases from the national surveillance system for notifiable infectious diseases [7]. The numbers of confirmed mpox cases were ordered according to date of symptom onset, throughout the analyses presented in this study. The earliest date of symptom onset among confirmed mpox cases in the Netherlands was 27 April 2022.

### Possible behavioural adaptations

We examined two possible adaptations: (1) MSM starting earlier to refrain from sexual contacts, thus resulting in shorter duration of the infectious period while not refraining from sex and (2) a decline in the number of casual partners [7]. We investigated scenarios with these behavioural adaptations taking place (a) only in July 2022 or (b) in June and in July 2022 [9, 10]. The magnitude and timing of the adaptations were obtained from the fitting process.

### Model fitting

The model was calibrated to the number of mpox cases using a Bayesian approach. We defined uniform prior distributions for the uncertain parameters (Table 1) and sampled 10,000 combinations of parameter values. In a first step, we started the model calculations with three infectious unvaccinated cases in the subgroup with very high sexual activity level. With each combination of parameter values, we computed the daily number of mpox cases and its Poisson likelihood up to 5 July 2022, for the scenarios with adaptations only in July 2022 (and up to17 June 2022, for the scenarios with adaptations in June and July 2022) – Table 3 shows parameters relating to the behavioural adaptations. This provided us the posterior distributions of all uncertain parameters except those relating to the behavioural adaptations that occurred in June/July 2022. In a second step, we used the posterior distributions obtained from the first step and fitted the model to data for the remaining days until 25 July 2022. The prior distributions of the parameters in the first and the second step are shown in Tables 1 and 3, respectively. The posterior distributions are shown in Tables S1-S3 in the Supplement.

**Table 3.** Prior distributions of uncertain parameters relating to the adaptations in behaviour of MSM that may have occurred in June/July 2022. Values were sampled from the uniform distribution with the ranges shown in this table.

|  | Days infectious<br>not refraining from<br>sexual contacts | Reduction* in number casual partners of group |  |  |
| --- | --- | --- | --- | --- |
|  |  | Low** | Fairly high | Very high |
| Scenario with adaptations at one time point: |  |  |  |  |
| After $T_2$ | 2-8 | 0-30% | 0-30% | 0-30% |
| Scenario with adaptations at two time points: |  |  |  |  |
| Between $T_1$ and $T_2$ | 2-8 | 0-30% | 0-30% | 0-30% |
| After $T_2$ | 2-8 | 0-30% | 0-30% | 0-30% |
| Dates when adaptations occurred: |  |  |  |  |
| $T_1$ | | 17-27 June 2022 | | |
| $T_2$ | | 5-15 July 2022 | | |

\* It was assumed that the number of casual partners of activity group $i$ was $\alpha_{ci}$ at the beginning of the outbreak (as estimated from the data – see Table 2) and was reduced to $(1 - D_{ij}) * \alpha_{ci}$ after day $T_j$ , with $D_{ij}$ sampled from the uniform distribution between 0 and 0.3. The maximum of 30% reduction was based on preliminary results from the survey “Monkeypox: a new challenge for your sex life” [9, 34] and reports from club owners in Amsterdam who experienced reduced numbers of visitors in July 2022, with the reduction reaching its maximum at the end of July 2022 with approximately 30% less visitors (P. Zantkuijl, personal communication).
\*\*Change applied to both very-low and fairly-low activity groups.

### Sensitivity analyses

Based on the size of the male population 17-66 years old [18] and available data on the prevalence of same-sex behaviour [19], we assumed that the number of MSM in 2022 was 250,000. Due to uncertainty in those estimates, we repeated the analyses with a population size of 200,000 or 300,000 MSM. Also, we carried out sensitivity analyses for the fraction of the population that had been vaccinated before 1975 via the old smallpox vaccination programme. In our main results, we assumed that 25% of the current MSM population had been vaccinated against smallpox in the past. We examined also two scenarios where (a) the percentage vaccinated in the past was higher among those with higher levels of sexual activity: 23%, 25%, 30%, 40% vaccinated in the group with very low, fairly low, fairly high, or very high level of sexual activity, respectively; and (b) the percentage vaccinated in the past was higher among those with lower levels of sexual activity: 29%, 25%, 15%, 10% vaccinated in the group with very low, fairly low, fairly high, or very high level of sexual activity, respectively. These percentages were chosen in such a way that in the overall MSM population 25% had obtained smallpox vaccination in the past.

### Hypothetical scenarios for the course of the outbreak without vaccination

Earlier work has indicated that the mpox outbreaks could fade out within a few months even without vaccination or other major interventions [4, 5, 12, 20]. To investigate this possibility, we examined hypothetical scenarios for the course of the mpox outbreak until the end of 2022, without the mpox vaccination programme.

We undertook model calculations until 31 December 2022 for the following scenarios: (a) without behavioural adaptations and (b) with behavioural adaptations in July 2022. For both scenarios, the parameter values were those obtained via the fitting process using data up to 25 July 2022. Also, we investigated whether new introductions of mpox cases could elicit a resurgence in mpox. For the scenario with behavioural adaptations in July 2022, we examined two scenarios with import of unvaccinated infectious mpox cases into the group with very high sexual activity level: (a) import of 20 cases on 1 November 2022 or (b) import of 1 case per day from 1 November until 31 December 2022.

## Results

### The decline in the mpox outbreak with and without behavioural adaptations

Without behavioural adaptations in the model, the daily number of mpox cases among MSM started declining around 8 July 2022, but the decline was less steep than observed (Figure 2a). The epidemic curve in the overall MSM population paralleled the epidemic curve in the group with very high sexual activity level (Figure 2b). Allowing for behavioural adaptations in July 2022, we found a decline of 13% (95% credible interval (CrI), 1-28%), 15% (95% CrI, 1-29%), and 24% (95% CrI, 1-30%) in numbers of casual partners of MSM with low, fairly high, and very high sexual activity, respectively, compared to before the mpox outbreak (Table S1 in Supplement). The infectious period while not refraining from sexual contacts was reduced from 6·0 days (95% CrI, 4·4-7·8 days) in May-June 2022 to 2·6 days (95% CrI, 2·0-4·3 days) in July 2022. The timing of these behavioural adaptations was calculated from the model at 7 July 2022 and there was a rather sharp decline in the number of cases thereafter (Table S1 and Figure 2c). With behavioural adaptations in June and July 2022, we found that the numbers of casual partners of MSM with low, fairly high, and very high sexual activity were reduced by 17%, 16%, and 18%, respectively, in June; and by 13%, 18%, and 22%, respectively, in July (Table S1). The infectious period while not refraining from sexual contacts was 6·9 days in May, 5·6 days in June, and 2·4 days in July 2022 (Table S1). The adaptations occurred as of 21 June and 10 July 2022 (Table S1 and Figure 2d).

**Figure 2.**
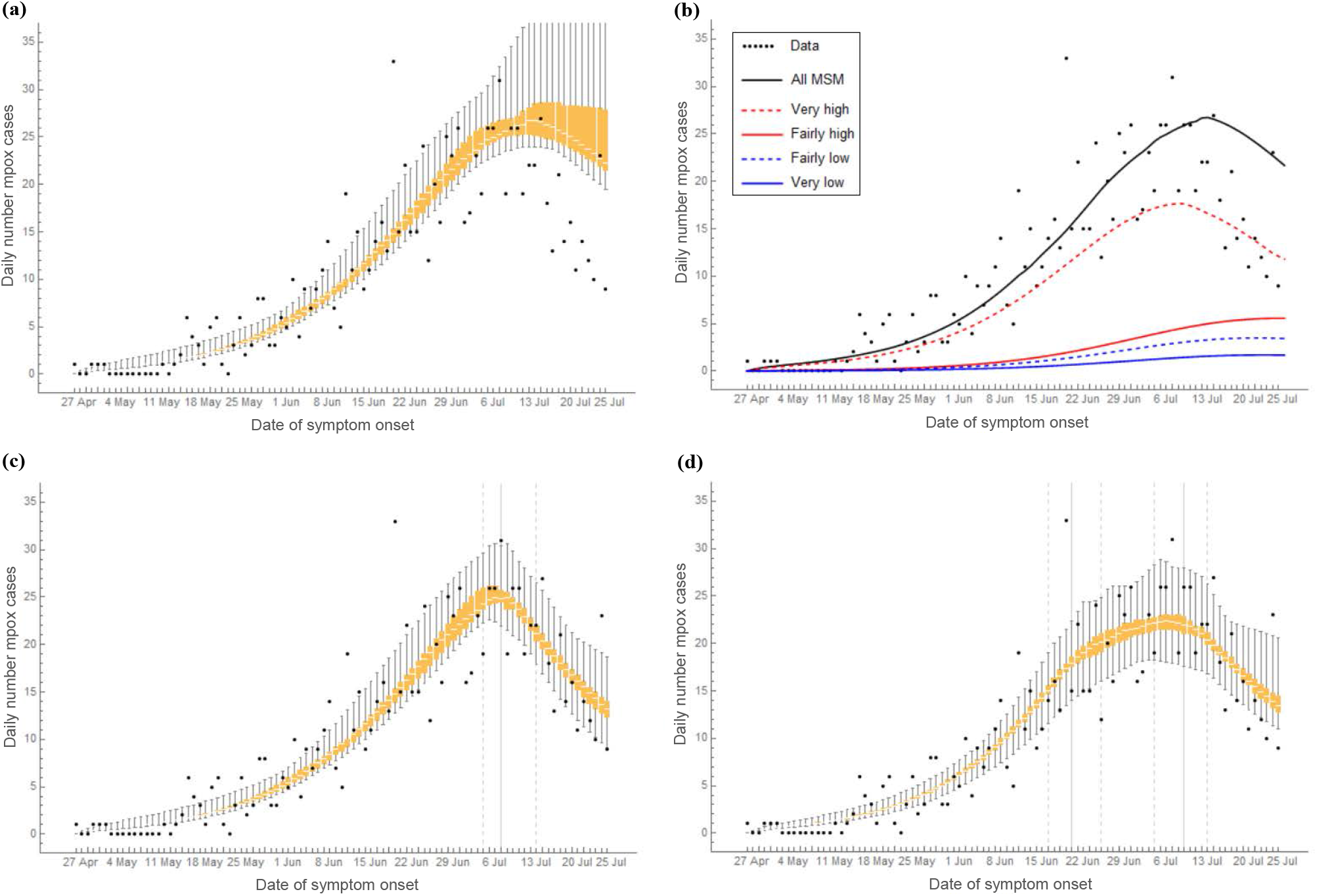
The daily number of mpox cases among MSM in the Netherlands from 27 April to 25 July 2022. Black bullets show data from the national database of notifiable diseases of the Netherlands. All other results were calculated from the model. Mpox cases are shown according to date of symptom onset. The orange box-plots show the daily numbers of mpox cases in the overall MSM population: (a) without behavioural adaptations; (c) with behavioural adaptations only in July 2022; (d) with behavioural adaptations in June and July 2022. (b) Median daily number of mpox cases in the overall MSM population (black line), in the groups with high sexual activity level (red lines), and in the groups with low sexual activity level (blue lines). In (c), (d), the vertical grey lines show medians (solid lines) and 95% credible intervals (dashed lines) of the day at which the behavioural adaptations occurred, as obtained from the model fitting. Model results were calculated in a population of 250,000 MSM.

### Distribution of mpox cases according to level of sexual activity

By 25 July 2022, 64% of mpox cases were MSM with very high and 18% with fairly high levels of sexual activity (Figure 3a,b). At that point, 57% (95% CrI, 33-66%) of the group with a very high sexual activity level had been infected with MPXV (Figure 3c), but only 1·1% (95% CrI, 0·9-1·3%) of the overall MSM population. Of all MPXV infections in the model until 25 July 2022, the infecting partner was a man with very high, fairly high, fairly low, or very low sexual activity in 76·7%, 18·7%, 3·9%, and 0·7% of infections, respectively (Figure 3d).

**Figure 3.**
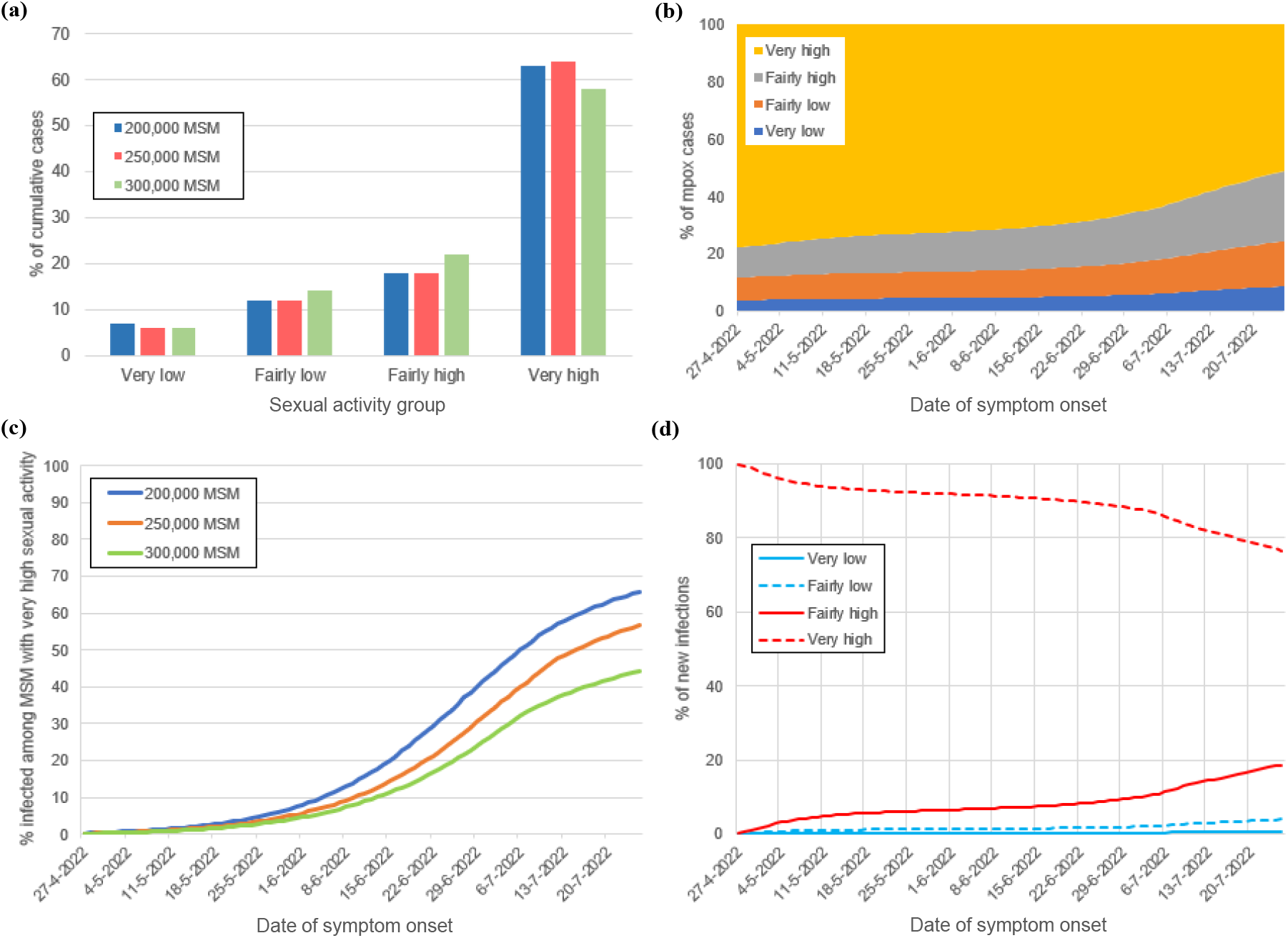
The role of subgroups of MSM with different levels of sexual activity, as calculated from the model, for the scenarios with behavioural adaptations in July 2022. (a) Distribution of cumulative mpox cases until 25 July 2022. (b) Distribution of daily mpox cases. (c) Percent of MSM with very high sexual activity being susceptible to monkeypox virus. (d) Distribution of new monkeypox virus infections among MSM according to the sexual activity group of the infecting individual. In (a), (c) results with population of 200,000; 250,000; or 300,000 MSM; in (b), (d) results with population of 250,000 MSM.

### Sensitivity analyses for the size of the MSM population

With a smaller population of 200,000 MSM (Figure S2, Table S2), the modelled decline in the daily number of cases started earlier and the number of cases in the very high sexual activity group was lower than in the scenario with 250,000 MSM. In a larger population of 300,000 MSM, the outbreak without behavioural adaptations would have been more severe than what we observed in the data (Figure S3, Table S2). The number of mpox cases in the model started declining around 20 July 2022, about ten days later than observed (Figure S3a). The fraction of the group with very high sexual activity level that had been infected with MPXV by 25 July 2022 was 66% in a population of 200,000 MSM and 45% in a population of 300,000 MSM (Figure 3c).

With different percentages of MSM vaccinated against smallpox in the past across the different sexual activity groups, the results were similar (Figure S4, Table S3). The transmission probability per sexual act was estimated at 0.60 (95% CrI, 0.47-0.80) with higher percentage vaccinated among those with higher numbers of sex partners (Table S3); 0.42 (95% CrI, 0.32-0.68), with higher percentage vaccinated among those with lower numbers of sex partners (Table S3); and 0.48 (95% CrI, 0.39-0.81), in the scenario with the same percentage vaccinated in all activity groups (Table S1).

### Hypothetical scenarios for the course of the outbreak without vaccination

Figure 4 shows the daily numbers of mpox cases from the model and from data until 31 December 2022. Without behavioural adaptations in 2022, the outbreak faded out by December 2022 with most of the parameter combinations (Figure 4a and S5). With the remaining parameter combinations, the modelled number of mpox cases reached its peak after July 2022, but it rapidly declined by the end of 2022. In 4.6% of the parameter combinations, we found more than 3 mpox cases by 31 December 2022 (Figure S5). These parameter combinations included high transmission probabilities combined with long infectious period without refraining from sexual contacts *and* low level of assortativeness in sexual mixing with casual partners. With behavioural adaptations in July 2022 in the model, the outbreak faded out by November 2022 (Figure 5b).

**Figure 4.**
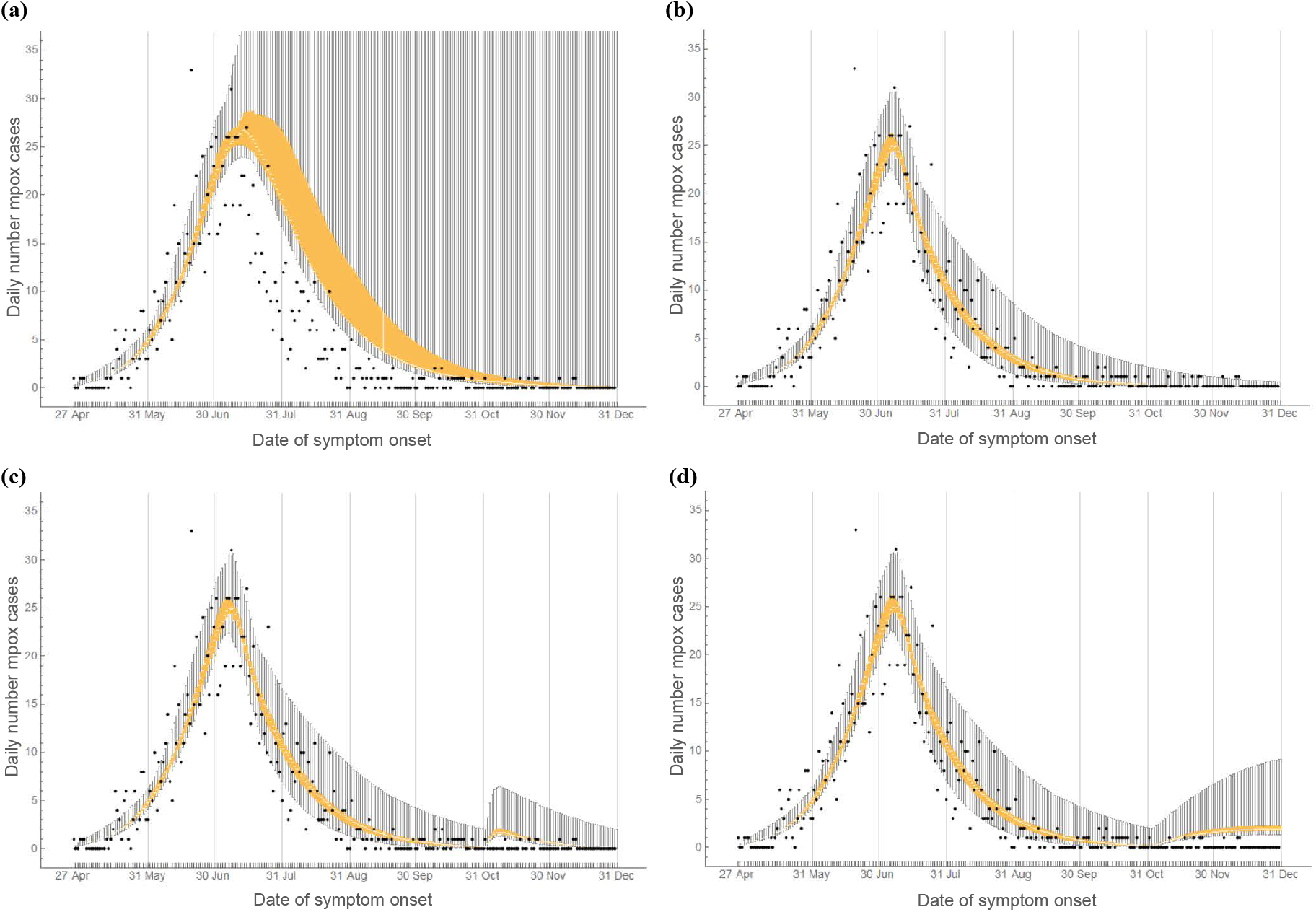
Daily number of mpox cases among MSM in the Netherlands from 27 April to 31 December 2022. Black bullets show data obtained from the national database of notifiable infectious diseases of the Netherlands. Orange box-plots show medians and interquartile ranges calculated from the model in a population of 250,000 MSM. Vertical solid grey lines show the last day of each calendar month. (a) Without behavioural adaptations. (b) With behavioural adaptations in July 2022. (c) With behavioural adaptations in July 2022 and import of 20 unvaccinated infectious mpox cases into the group with very high level of sexual activity on 1 November 2022. (d) With behavioural adaptations in July 2022 and import of one infectious case per day (in the group with very high level of sexual activity) from 1 November to 31 December 2022.

Import of unvaccinated infectious MSM with very high level of sexual activity resulted in a small increase in mpox cases. With import of 20 cases on 1 November 2022, we found 41 (95% CrI, 30-106) additional mpox cases (Figure 4c); with import of one case per day from 1 November to 31 December 2022, we found 81 (95% CrI, 63-211) additional cases (Figure 4d), compared to the scenario without import.

## Discussion

We developed a transmission model of the mpox outbreak among MSM and fitted this model to the daily numbers of mpox notifications in the Netherlands. We demonstrated that depletion of susceptibles among MSM with very high levels of sexual activity may have limited further growth of the mpox epidemic, but could not solely account for the rapid decline after the peak. This suggests that behavioural adaptations may have accelerated the decrease in mpox cases. Furthermore, our findings suggest that, even without the mpox vaccination programme, the level of immunity due to infection among MSM with a very high level of sexual activity was sufficiently high to lead to the fading out of the outbreak by November 2022.

The major strength of our study is that we quantified the contribution of infection-induced immunity and the contribution of behavioural adaptations in shaping the outbreak. The model, informed by the daily numbers of registered mpox cases during the rise and the decline of the mpox outbreak, allowed us to explore counterfactual situations and to show that the outbreak could have waned by the end of 2022 even without mpox vaccination or behavioural adaptations.

We incorporated heterogeneity in sexual activity by dividing the population into groups with different levels of sexual activity and captured the assortative mixing of highly active individuals that is characteristic of sexual contact networks. This modelling approach has the advantage of being well established, validated, and extensively studied in the past four decades in the context of several sexually transmitted infections [21-24]. Heterogeneity in the number of sexual partners and assortative mixing may enhance the selective spread of infection among individuals with higher sexual activity (see [21, 22] and references herein). Our findings are consistent with previous studies on MPXV transmission showing that a very small group of MSM with high level of sexual activity can be sufficient to cause large outbreaks of short duration [4, 12, 20]. Our results correspond well also with a study that showed how the observed decline in mpox cases could be primarily attributed to infection-induced immunity [5]. The present study extends earlier findings as our results quantify the contribution of infection-induced immunity and the contribution of behavioural adaptations and reveal how these factors shaped the outbreak. In contrast with other studies projecting smaller outbreaks [25] or more severe and longer-lasting outbreaks [4, 12, 13], we found that the size of the 2022 mpox outbreak was large but self-limited and could have died out within a few months even without behavioural adaptations or vaccination.

We showed that, without behavioural adaptations, the decline in mpox cases in a larger population of 300,000 MSM started later than what we observed in the data. This suggests that the observed decline could have been possible only with behavioural adaptations. When extending the model results without behavioural adaptations to the end of 2022, we found that in a small fraction of the fitted parameter combinations, the number of mpox cases peaked after July 2022 and the outbreaks faded out after the end of 2022. In this situation, behavioural adaptations and vaccination could have prevented a large number of cases.

Our results suggest that a reduction in numbers of sexual partners considerably expedited the decline in mpox cases. MSM may have started changing their behaviour with the first news of mpox in the general media. This effect may have been strengthened by the mpox awareness campaigns that were conducted in gay sex venues and media [10, 26]. Accurate and timely communication with populations most at risk appears essential for public health response, also as similar outbreaks could occur with other pathogens when introduced into subpopulations with sufficiently high contact levels. The mpox vaccination programme possibly accelerated the impact of awareness and behavioural adaptations and offered protection against mpox at the individual level. Vaccination also reduced the risk of resurgence of mpox, since it enhanced immunity at the population level. Our model can be used to investigate the impact of vaccination for specific subgroups of MSM. Furthermore, our model could be extended to account for transitioning across sexual activity groups.

Our study has important implications for the prevention of mpox outbreaks in the future. Prevention can be achieved by maintaining a high level of immunity among those with the highest sexual activity level. First, it should be taken into account that the group of MSM with the highest sexual activity level may not be a closed and fixed group over time: it is likely that over long time periods, some turnover occurs among this group and the rest of the MSM population. For example, MSM with low levels of sexual activity who did not receive vaccination in 2022 might become more sexually active and replenish the pool of susceptible MSM with high level of sexual activity [22], thus making a new mpox outbreak possible.

Second, in assessing the effectiveness of public health messages and interventions, it is important to consider the venues that the target group might visit. Having sex in sex clubs and parties has been identified as an independent predictor of MPXV infection [9]. Third, in the current study, we assumed that the percentage of MSM that had received smallpox vaccination in the past was the same in each sexual activity group. Our sensitivity analysis indicates that this assumption had a limited impact on the results. However, the percentages immune due to the old smallpox vaccination programmes may not be exactly the same across the MSM population, depending on the age distribution of men in each sexual activity group. For future prevention of mpox outbreaks it is important to include the changing distribution of immunity with age.

Caution is needed when extrapolating our results to other settings. We allowed for only small behavioural adaptations. With the high transmission potential of MPXV, it is possible that with less assortative sexual mixing, the virus could have spread faster among the groups of MSM with low sexual activity levels. In that case, infection-induced immunity within the group with very high sexual activity level might have increased slower than in our analyses, making persistence more likely. Then fading of the outbreak would not have been possible without large reductions in numbers of sex partners. However, this is unlikely for the mpox outbreak in the Netherlands, since monthly numbers of STI diagnoses, tests, and consultations were more or less stable throughout 2022 [27], therefore, we have no evidence of a large decline in numbers of sexual partners of MSM. Finally, recent reports point to the possibility of reinfections [28], but this was not accounted for in this study.

In conclusion, we demonstrated that depletion of susceptible MSM with high levels of sexual activity as well as behavioural adaptations accounted for the timing and speed of the decline in mpox cases among MSM in the Netherlands. The level of immunity due to infection within the subgroup of MSM with the highest level of sexual activity was build up so fast that the outbreak could have waned within a few months, even without interventions or behavioural adaptations. The risk of a resurgence of mpox in the short term seems very low, even with import of mpox infections.

## Data Availability

The data for the numbers of daily mpox cases in the Netherlands described in this study can be freely and openly accessed on the website of the National Institute of Public Health and the Environment (RIVM) of the Netherlands: https://www.rivm.nl/en/mpox. All other data (used as input to inform model parameters; and data obtained as model output) are available upon reasonable request to the authors

## Acknowledgements

The authors would like to thank Paul Zantkuijl (Soa Aids Nederland) for information on mpox messages in media and health promotion platforms for MSM in the Netherlands and for information about the reduction in visitors in clubs and sex venues in Amsterdam in July 2022. Anonymous club owners are also thanked for providing the above information. We express our gratitude to the participants, the research teams, and the organizations involved in the “COVID-19, Sex, and Intimacy Survey” and the survey “Monkeypox: a new challenge for your sex life” led by Philippe Adam and John de Wit. Both surveys were conducted in partnership between Utrecht University, the Institute for Prevention and Social Research (IPSR Utrecht), Soa Aids Nederland, and the National Institute of Public Health and Environment (RIVM). We thank the anonymous reviewers for providing comments and suggestions that substantially improved the paper.

## SUPPLEMENT

### The transmission model

We developed a deterministic compartmental model that describes monkeypox virus (MPXV) transmission among men who have sex with men (MSM). A schematic diagram of the model is shown in Figure 1 in the main text. We accounted only for transmission via sexual or intimate contacts among main (steady) and casual sex partners (for brevity, referred to as “sexual contacts”).

### Sexual partners

MSM were divided into *G* = 4 sexual activity groups, based on the total number of partners with whom they had sexual/intimate contacts. The number of these partners was obtained from data from the first round of the “COVID-19, Sex, and Intimacy Survey”, carried out in the summer of 2020 [1]. The survey was focussed on the impact of the first wave of the pandemic. However, as reference, participants were asked also about their sexual behaviour in the second half of 2019. We used the data of 2019, as representative of sexual activity without the temporal fluctuations caused by the COVID-19 pandemic, but also because participants were asked to report the number of male partners with whom they “had sex/intimacy in the second half of 2019”. Based on this number, we divided the population into four sexual activity groups: very low, fairly low, fairly high, and very high activity. The parameters relating to sexual behaviour of each activity group are shown in Table 2.

### The course of MPXV infection

Individuals enter the population as susceptible (*S*_*ui*_) when they become sexually active. After infection, individuals are exposed but not infectious (*E*_*ui*_) and later become infectious (*I*_*ui*_). When individuals have symptoms and/or they are tested positive for MPXV infection, they may refrain from physical/sexual contacts or may be in isolation/confinement. We refer to these individuals as “refraining from sexual contacts” (*Y*_*ui*_). We assumed that individuals start refraining from sexual contacts 1/δ days after becoming infectious; infectious cases refraining from sexual contacts can still infect others, with a lower probability of transmission, for instance, because they do not completely refrain from sex contacts. Most of the infectious mpox cases recover after a few weeks and become immune (*R*_*i*_), but a small fraction of the infectious cases may develop complications and need hospitalization (*H*_*i*)_ or die due to mpox.

Individuals born before 1974 were vaccinated in their first year of age against smallpox and that protects against MPXV infection and/or disease. Smallpox vaccination ended in 1974 in the Netherlands. Therefore, we assumed that in 2022 approximately 25% of sexually active MSM is vaccinated (*S*_*Vi*_) via the old smallpox vaccination programmes, based on the age distribution of adult men in the Netherlands [2, 3]. We assumed that vaccinated men can still get infected with MPXV (and become exposed, *E*_*Vi*_), but at a lower rate than that for unvaccinated individuals. Vaccinated exposed individuals have a lower rate of becoming infectious (*I*_*Vi*_), than unvaccinated exposed individuals. Vaccinated infectious cases may refrain from sexual contacts (*Y*_*Vi*_). We assumed that vaccination reduces the level of infectivity of vaccinated infectious cases, but the recovery rate, the hospitalization rate, and the MPXV-related death rate are similar for vaccinated and unvaccinated individuals.

To account for different levels of protection by the old and the new vaccines, in the model we included individuals vaccinated in 2022 separately from those who had been vaccinated in the past (before 1974). The respective compartments in the model are denoted with *S*_*pi*_, *E*_*pi*_, *I*_*pi*_, *Y*_*pi*_ (uninfected, exposed, infectious not refraining from sexual contacts, infectious refraining from sexual contacts, respectively) with the subscript *p* denoting those vaccinated in 2022 by means of pre-exposure or post-exposure prophylaxis. Susceptible individuals (unvaccinated or vaccinated in the past) may be vaccinated in 2022 at a rate *φ*_1*i*_. Exposed individuals (unvaccinated or vaccinated in the past) may be vaccinated in 2022 at a rate *φ*_2*i*_. The vaccination rates *φ*_1*i*_ and *φ*_2*i*_ depend on the activity group *i*. We assumed that the protection of the new vaccine (against infection and disease) is higher than that from the old vaccine (Table 1). Both vaccine protections were modelled as a proportional reduction in the transmission rate.

Hospitalized cases may die due to mpox or recover and become immune (*R*_*i*_). All MSM entering the sexually active population in 2022 are unvaccinated and susceptible to MPXV.

### Model equations

The model is described by the system of ordinary differential equations shown below. The subscript *i* denotes unvaccinated individuals, *V* denotes individuals vaccinated in the past (with the old vaccine), *p* denotes individuals vaccinated in 2022 (with the new vaccine), and *i* = 1,2, …, *G* denotes the *i*-th sexual activity group.

Equations for unvaccinated individuals:

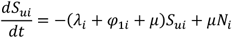

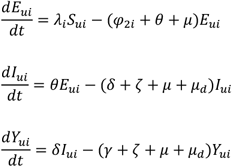

Equations for individuals vaccinated in the past:

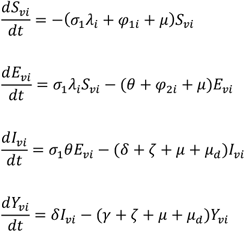

Equations for individuals vaccinated in 2022:

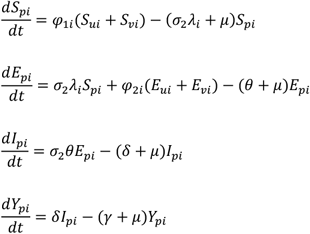

Equations for recovered/immune and hospitalized individuals:

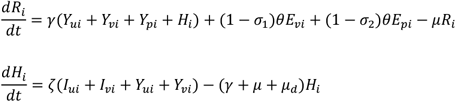

The parameters and variables in these equations are explained in the following sections and in Tables 1-3.

### Transmission rate

The rate at which MSM in activity group *i* get infected with MPXV is λ_*i*_ = λ_*mi*_ + λ_*ci*_, where λ_*mi*_ and λ_*ci*_ denote the rates of getting infected by main and casual partners, respectively:

- The rate of getting infected by main regular partners:

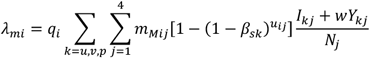
- The rate of getting infected by casual partners:

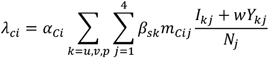

In these equations, the following notation is used:

- *q*_*i*_ is the fraction of MSM of activity group *i* with a main regular partner.
- *ui*_*ij*_ is the frequency of sex contacts between main sexual partners of activity groups *i, j*, calculated as *ui*_*ij*_ = (*ui* + *i*_*ij*_)/2, from the contact frequency *ui* and *i*_*ij*_ of sexual activity groups *i* and *j*, respectively.
- β_*sk*_ is the probability of transmission of MPXV per sexual/intimate contact from an infectious individual who is unvaccinated (*k* = *i*), vaccinated in the past (*k* = *V*), or vaccinated in 2022 (*k* = *p*), with β_*su*_ = β_*su*_, β_*sv*_ = *V*_1_β_*s*_, β_*sp*_ = *V*_2_β_*s*_. The probability β_*s*_ of transmission per sexual contact with an unvaccinated individual is reduced by *V*_1_ for those who were vaccinated in the past and by *V*_2_ for those vaccinated in 2022.
- *W* is a factor reducing the transmission potential of an infectious individual practicing sexual abstinence compared to infectious individuals not in abstinence.
- *α*_*Ci*_ is the number of casual sex contacts per day for men in activity group *i*.
- *N*_*ij*_ is the total size of activity group *j* and 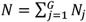 is the total size of the MSM population.
- *m*_*mij*_ and *m*_*Cij*_ are parameters that define the level of mixing between sexual activity groups *i, j*, when forming main and casual partnerships, respectively. These are defined by the equations:

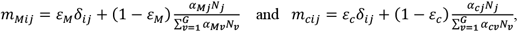

where δ_*ij*_ is the Kronecker delta (being equal to 1, if *i* = *j*; and equal to 0, otherwise) and the parameters *ε*_*s*_, *ε*_*c*_ determine the level of assortativeness in mixing of activity groups when forming main and casual partnerships, respectively (if *ε*_*i*_ = 1, then mixing is assortative; if *ε*_*i*_ = 0, then mixing is proportionate).

### The size of the MSM population

Estimates of the size of the MSM population vary from 111,072 [4] to 392,000 [5]. The variation depends mostly on the ages included in the estimate (for instance, from 15 or 17 years old; up to 65 or 69 years old) and the definition of MSM (men who had sex with men in the previous six/twelve months; or ever having sex with men; or identifying themselves as homosexual/bisexual). Based on the size of the male population [2] and prevalence estimates of same-sex behaviour [5], we assumed that the number of MSM in 2022 was around 250,000. Due to uncertainty about this estimate, we repeated the analyses with 200,000 MSM and 300,000 MSM.

### Adaptations that may have occurred in the first three months of the outbreak

By ordering diagnosed mpox cases according to date of symptom onset, the peak was between 6 and 10 July 2022. The first diagnosis of mpox was on 20 May 2022. By the beginning of June, the number of diagnoses was increasing and there were messages about mpox in the news, social media, and MSM websites [6-9]. This has probably enhanced awareness among MSM and health care practitioners, enabling earlier recognition of mpox symptoms, even during the prodromal phase with systemic symptoms like fever, fatigue, and ache. In an online survey carried out in August 2022, most MSM responded correctly to questions about symptomatology and routes of transmission of mpox and reported to be willing to refrain from close physical and sexual contacts if they were infected with MPXV [10]. As the severity of the outbreak was increasing, MSM were possibly more willing to refrain from sexual/intimate contacts when they suspected or were diagnosed with MPXV infection, thus reducing the time they were infectious and not yet refraining from sexual contacts. The spread of information about mpox and the severity of the outbreak may have also influenced the sexual behaviour of MSM. In the same survey, more than half of the participants reported a reduction in their sexual activity due to the mpox outbreak [10]. Furthermore, sex venues and parties reported low numbers of visitors in July 2022 [11], with the reduction reaching its maximum at the end of July, when some club owners observed up to 30% less visitors than what was expected (P. Zantkuijl, personal communication). This points to a decline in sexual activity of men visiting these accommodations.

Therefore, in this study, we examined two possible adaptations: (a) MSM may start refraining from sexual contacts earlier during the infectious period, thus resulting in shorter infectious period while not refraining from sexual contacts and (b) a reduction in the number of casual partners. The level and the timing of the adaptations was obtained from the fitting process. We examined two scenarios with adaptations either (a) at one time point *T*_2_ in the period 5-15 July 2022 or (b) at two time points *T*_1_ in the period 17-27 June 2022 and *T*_2_ in the period 5-15 July 2022. These time periods were close to messages placed on ManTotMan socials and the announcement of the Dutch government on 7 July 2022 to start with the monkeypox vaccination programme.

We included a reduction *D*_4*j*_ in the number of casual partners of men in the very high sexual activity group, *D*_3*j*_ for men in the fairly high sexual activity group, and a reduction *D*_2*j*_ for men in the very low and fairly low sexual activity groups. The reduction *D*_*ij*_ occurred on day *T*_*j*_ = *T*_1_ or *T*_*j*_ = *T*_2_. The six values *D*_*ij*_ (*i* = 2,3,4 and *j* = 1,2) were sampled from the same range (0-30% reduction), since we are uncertain about the occurrence of the decline. Similarly, we sampled three values for the duration of the infectious period before refraining from sexual contacts: one for the duration in the beginning of the outbreak, a second value for the period between *T*_1_ and *T*_2_, and a third value for the period after *T*_2_. All three values were sampled from the same range (2-8 days) since we were uncertain whether an adaptation in this duration occurred and when.

### Model fitting

The model was fitted to data on the number of diagnosed mpox cases using a Bayesian approach. Parameters relating to sexual behaviour were estimated from data or obtained from the literature (Table 2), except from: (a) the two parameters for assortativeness in sexual mixing and (b) the number of casual partners of the group with a very high sexual activity level. These were included as uncertain parameters, due to lack of reliable data and because the model results were very sensitive to these parameters (based on our preliminary analyses). Further, most of the parameters relating to mpox were uncertain (Table 1) and were obtained via the fitting process.

The uncertain parameters were divided into two groups:

1. Parameters relating to behavioural adaptations that occurred in June/July 2022 (Table 3): adaptation in the number of casual partners; adaptation in the number of days that an individual is infectious and not in abstinence; and two parameters for the timing of these adaptations.
2. Main parameters (Table 1): all the other parameters, except those relating to the adaptations that occurred in June/July 2022.

The fitting process was carried out in two steps:

1. In the first step, we fitted the model to the numbers of daily mpox cases registered in the national database of notifiable infectious diseases of the Netherlands until 17 June or until 5 July 2022. We defined uniform prior distributions for the uncertain parameters (Tables 1). Using Latin Hypercube Sampling [12], we sampled 10,000 combinations of values from the prior distributions and repeated the model calculations with each parameter combination. From the model, we calculated the daily numbers of mpox cases with each parameter combination. We calculated the Poisson likelihood of the above numbers, thus obtaining the posterior distributions of the uncertain parameters (except those relating to the behavioural adaptations in June/July 2022), reflecting the situation in the beginning of the outbreak.
2. In the second step, we used the posterior distributions obtained from the first step of the fitting process and fitted the model to data from 17 June or 5 July 2022 until 25 July 2022. The modelled mpox cases were compared with the respective data with date of symptom onset in this time interval. We calculated their Poisson likelihood and obtained the posterior distributions of the uncertain parameters relating to behavioural adaptations that occurred in June/July 2022.

### Data sources

The following data sources were used in this study.

#### National surveillance system for notifiable diseases (OSIRIS)

Individuals seeking mpox testing or presenting with symptoms suggestive of mpox to general practitioners or health centra were referred and notified to the regional public health service for mpox diagnostics [9]. Notification of confirmed mpox cases from regional public health services to the National Institute of Public Health and the Environment was accompanied by a questionnaire with demographical, clinical, and epidemiological information of cases. The numbers of confirmed mpox cases according to date of symptom onset were extracted from this database. These numbers were continuously updated during the outbreak and were available online on https://www.rivm.nl/mpox-apenpokken.

#### COVID-19, Sex, and Intimacy Survey

This is a repeated cross-sectional self-report survey. The first round of data collection was carried out from the end of July 2020 to the beginning of September 2020 [13, 14].

Participants were recruited via social media advertisement on Facebook and Instagram. People were eligible to participate if they: lived in the Netherlands, were 18 years or older, identified as male and ever had sex with a man. All participants provided informed consent and received no compensation. Only responses from eligible participants who fully completed the questionnaire were used for the analyses presented here. The aim of the survey was to investigate the impact of the COVID-19 pandemic on sexual behaviour. Therefore, respondents were asked to report on their sexual behaviour during the first half of 2020 and, as reference, their sexual behaviour during the last six months of 2019. In the present study, we used data only from the responses for the period July-December 2019. For these questions, 5,683 participants reported their sexual behaviour.

**Figure S1.**
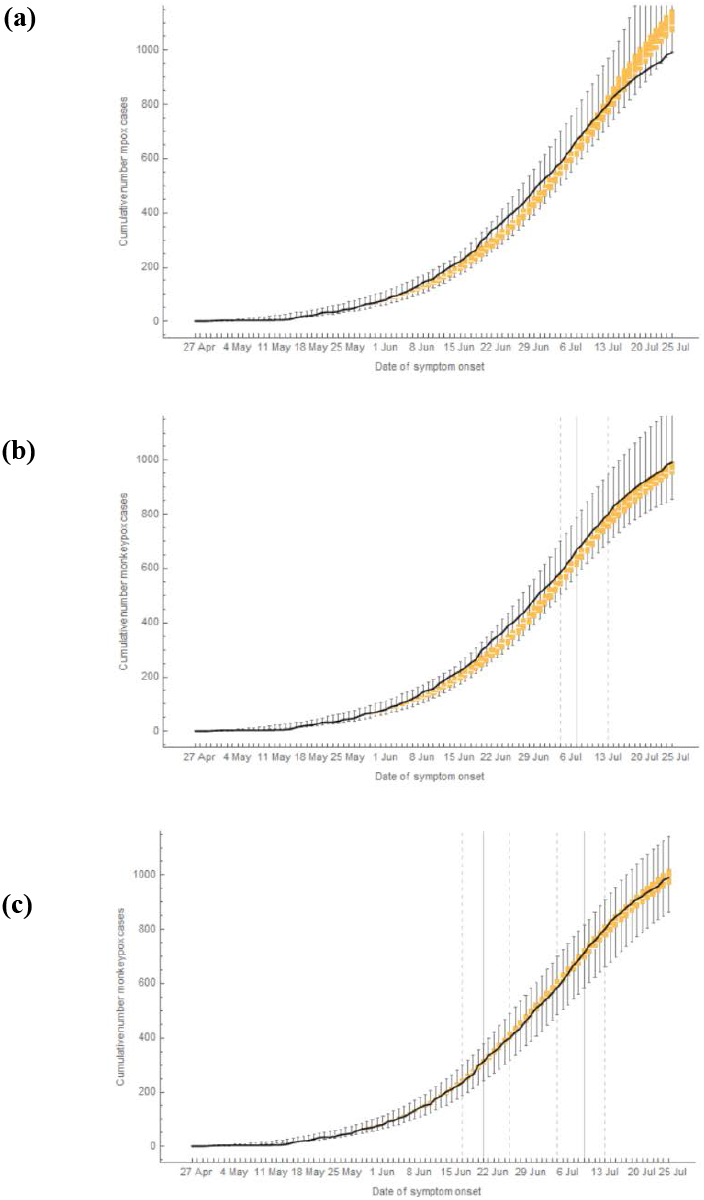
The cumulative number of mpox cases among MSM in the Netherlands from 27 April to 25 July 2022. Cases are shown according to date of symptom onset (on horizontal axes). Solid black line shows data from 27 April to 25 July 2022, from the national database of notifiable infectious diseases of the Netherlands. Orange box-plots show the medians and interquartile ranges obtained from the model in a population of 250,000 MSM (a) without behavioural adaptations; (b) with behavioural adaptations only in July 2022; (c) with behavioural adaptations in June and in July 2022. In (b)-(c), the vertical grey lines show the medians (solid line) and the 95% credible intervals (dashed lines) of the time point at which the adaptations occurred, as obtained from the model fitting.

**Figure S2.**
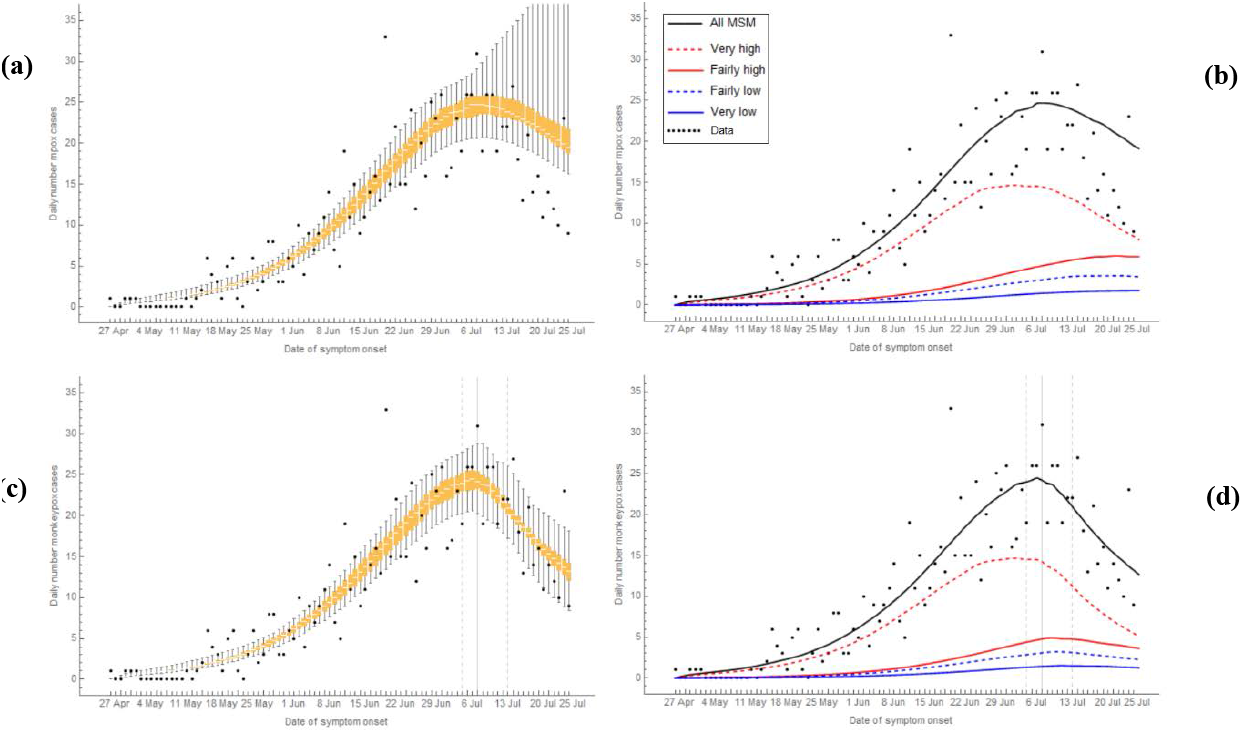
The daily number of mpox cases among MSM in the Netherlands from 27 April to 25 July 2022. Black bullets show data from the national database of notifiable diseases of the Netherlands; the other lines were calculated from the model. Mpox cases are shown according to date of symptom onset. Left panels: box-plots of daily number of mpox cases in the overall MSM population. Right panels: median daily number of mpox cases in the overall MSM population (black line), in the groups with high sexual activity level (red lines), and in the groups with low sexual activity level (blue lines). (a), (b): without behavioural adaptations; (c), (d): with behavioural adaptations in July 2022. Vertical grey lines show median (solid line) and 95% credible interval (dashed lines) of the day at which the behavioural adaptations occurred, as obtained from the model fitting. Model results were calculated in a population of 200,000 MSM.

**Figure S3.**
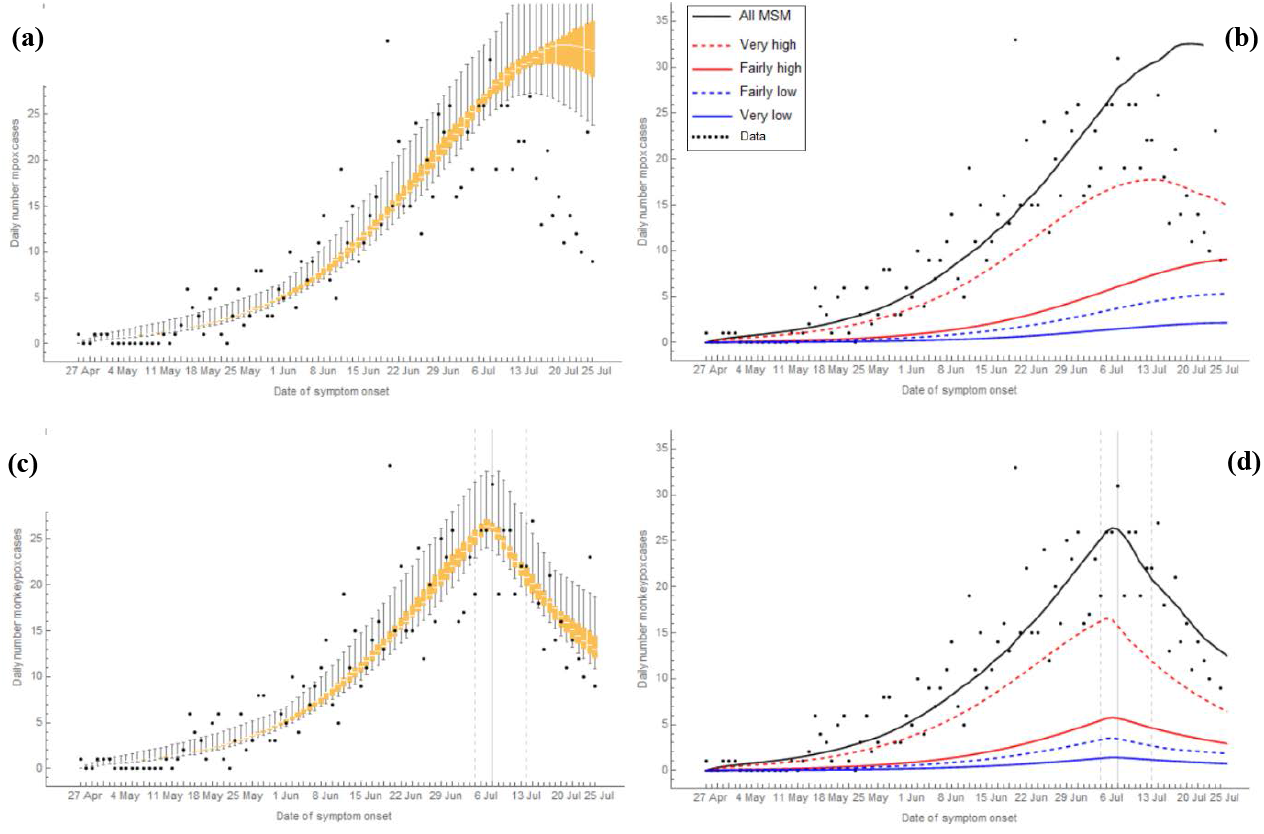
The daily number of mpox cases among MSM in the Netherlands from 27 April to 25 July 2022. Black bullets show data from the national database of notifiable diseases of the Netherlands; the other lines were calculated from the model. Mpox cases are shown according to date of symptom onset. Left panels: box-plots of daily number of mpox cases in the overall MSM population. Right panels: median daily number of mpox cases in the overall MSM population (black line), in the groups with high sexual activity level (red lines), and in the groups with low sexual activity level (blue lines). (a), (b): without behavioural adaptations; (c), (d): with behavioural adaptations in July 2022. Vertical grey lines show median (solid line) and 95% credible interval (dashed lines) of the day at which the behavioural adaptations occurred, as obtained from the model fitting. Model results were calculated in a population of 300,000 MSM.

**Figure S4.**
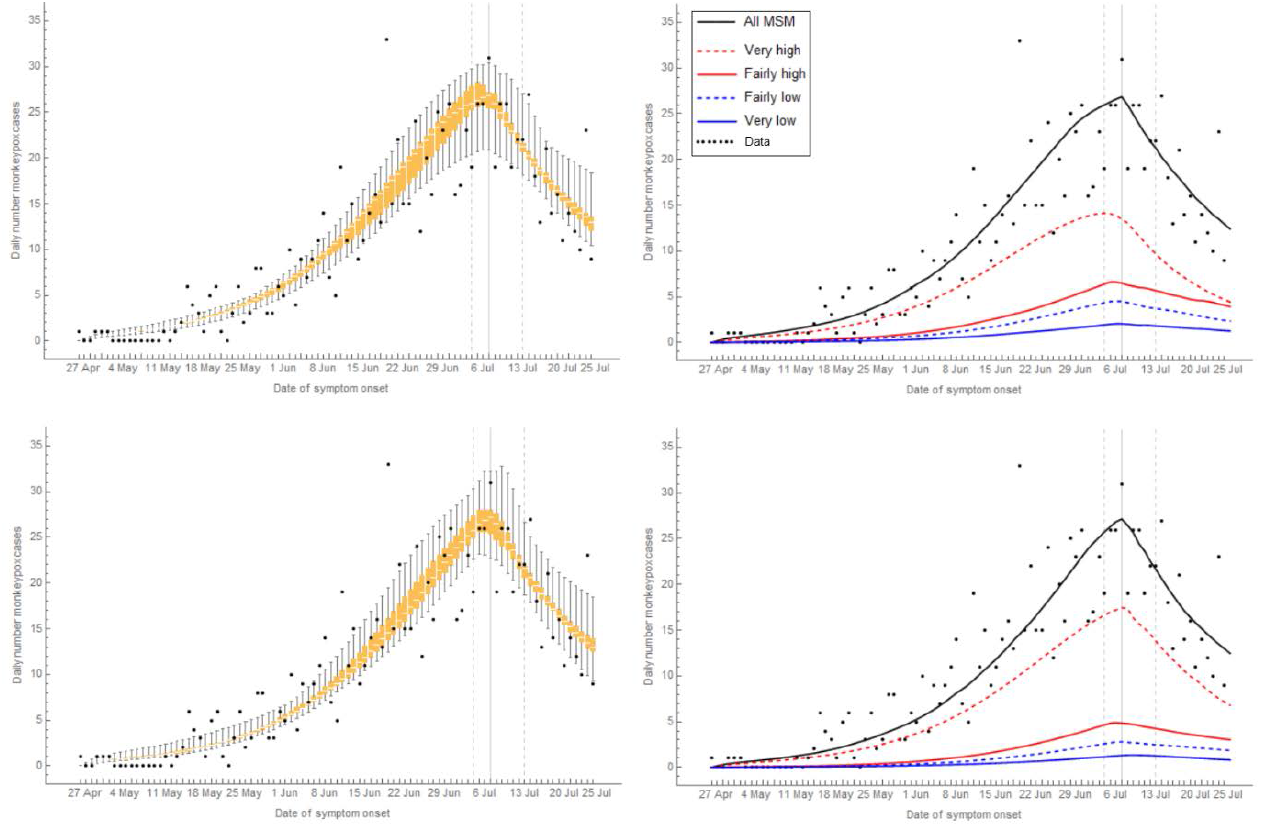
Sensitivity analyses for the percentage of the population being vaccinated in the past (before 1974), via the old smallpox vaccination programme. Two scenarios are shown, with behavioural adaptations in July 2022, in a population of 250,000 MSM, of whom 25% in total had been vaccinated in the past. Top panels: the percentage vaccinated in the past was the highest in the group with the highest level of sexual activity: 23%, 25%, 30%, 40% vaccinated in the group with very low, fairly low, fairly high, or very high level of sexual activity, respectively. Bottom panels: the percentage vaccinated in the past was the highest in the group with the lowest level of sexual activity: 29%, 25%, 15%, 10% vaccinated in the group with very low, fairly low, fairly high, or very high level of sexual activity, respectively. Numbers shown are the daily numbers of mpox cases among MSM in the Netherlands from 27 April to 25 July 2022, from the data (black bullets) and from the model (orange box plots and lines) – the plots are as explained in Figure S3.

**Figure S5.**
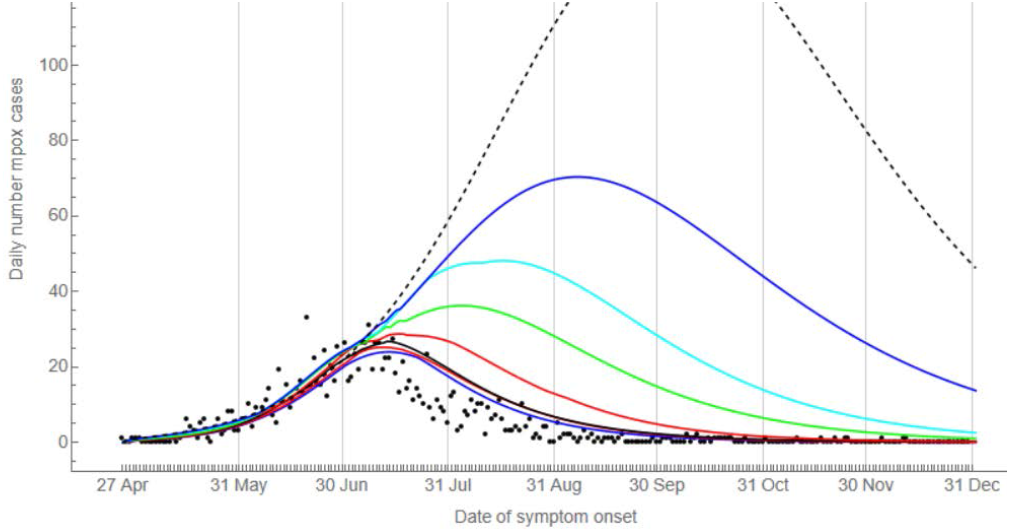
The daily number of mpox cases among MSM in the Netherlands from 27 April to 31 December 2022, in the scenario without behavioural adaptations. Black bullets show data from the national database of notifiable diseases of the Netherlands, as shown in Figure 4a in the main text. The lines were calculated from the model. Mpox cases are shown according to date of symptom onset. Black solid line: median; black dashed line: maximum; red lines: interquartile range; blue lines, 95% credible interval; cyan line, upper 5^th^ percentile; green line, upper 10^th^ percentile. Vertical grey lines show the last day of each calendar month. Model results were calculated in a population of 250,000 MSM.

**Table S1.** Posterior distributions of uncertain model parameters obtained from the fitting process for the two scenarios with adaptations in behaviour of MSM only in July 2022 or in June and July 2022. Results shown are the medians and 95% credible intervals, in a population of 250,000 MSM.

|  | Adaptations only in July 2022 |  |  | Adaptations in June & July 2022 |  |  |
| --- | --- | --- | --- | --- | --- | --- |
|  | Median | 95% range |  | Median | 95% range |  |
| Latent period, days ($1/\theta$) | 5.67 | 5.03 | 6.97 | 5.80 | 5.10 | 7.81 |
| Infectious period ($1/\gamma + 1/\delta$) | 16.75 | 14.27 | 24.07 | 18.41 | 14.26 | 24.46 |
| Transmission probability per sex act | 0.48 | 0.39 | 0.81 | 0.48 | 0.39 | 0.67 |
| Number casual partners per day, very-high activity group | 0.89 | 0.73 | 1.00 | 0.90 | 0.73 | 1.00 |
| Factor reducing transmission when in abstinence ($w$) | 19% | 15% | 37% | 22% | 16% | 46% |
| Assortativeness mixing with main partners ($\varepsilon_M$) | 0.72 | 0.13 | 0.88 | 0.69 | 0.12 | 0.85 |
| Assortativeness mixing with casual partners ($\varepsilon_C$) | 0.87 | 0.20 | 0.90 | 0.84 | 0.48 | 0.90 |
| Vaccination rate before 25 July 2022 | 0.04% | 0.03% | 0.05% | 0.04% | 0.03% | 0.05% |
| Efficacy old vaccine ($v_1 = \sigma_1$) | 56% | 52% | 65% | 58% | 50% | 63% |
| Efficacy new vaccine ($v_2 = \sigma_2$) | 85% | 80% | 90% | 85% | 80% | 89% |
| Hospitalization rate, $\zeta$ | 1% | 1% | 2% | 2% | 1% | 2% |
| Days infectious not refraining from sex, before $T_1$ | 6.05 | 4.45 | 7.80 | 6.89 | 3.74 | 7.78 |
| Days infectious not refraining from sex, between $T_1$ and $T_2$ | As before $T_1$ | | | 5.60 | 2.94 | 7.83 |
| Days infectious not refraining from sex, after $T_2$ | 2.55 | 2.00 | 4.31 | 2.44 | 2.02 | 4.87 |
| Date of adaptations in June, $T_1$ | Not applicable | | | 21-6-2022 | 17-6-2022 | 26-6-2022 |
| Date of adaptations in July, $T_2$ | 7-7-2022 | 5-7-2022 | 11-7-2022 | 10-7-2022 | 5-7-2022 | 14-7-2022 |
| % reduction casual partners, low activity groups, after $T_2$ | 13% | 1% | 28% | 13% | 0% | 28% |
| % reduction casual partners, fairly-high activity group, after $T_2$ | 15% | 1% | 29% | 18% | 3% | 29% |
| % reduction casual partners, very-high activity group, after $T_2$ | 24% | 1% | 30% | 22% | 4% | 30% |
| % reduction casual partners, low activity groups, between $T_1$ and $T_2$ | Not applicable | | | 17% | 2% | 29% |
| % reduction casual partners, fairly-high activity group, between $T_1$ and $T_2$ | | | | 16% | 1% | 29% |
| % reduction casual partners, very-high activity group, between $T_1$ and $T_2$ | | | | 18% | 1% | 29% |

**Table S2.** Posterior distributions of uncertain model parameters obtained from the fitting process for the scenario with adaptations in behaviour of MSM only in July 2022. Results shown are the medians and 95% credible intervals, in a population of 200,000 or 300,000 MSM.

|  | 200,000 MSM |  |  | 300,000 MSM |  |  |
| --- | --- | --- | --- | --- | --- | --- |
|  | Median | 95% range |  | Median | 95% range |  |
| Latent period, days ($1/\theta$) | 5.51 | 5.04 | 7.77 | 5.67 | 5.04 | 7.74 |
| Infectious period ($1/\gamma + 1/\delta$) | 18.41 | 14.51 | 26.75 | 15.54 | 14.22 | 25.88 |
| Transmission probability per sex act | 0.50 | 0.39 | 0.66 | 0.49 | 0.45 | 0.81 |
| Number casual partners per day, very-high activity group | 0.91 | 0.72 | 1.00 | 0.90 | 0.72 | 0.98 |
| Factor reducing transmission when in abstinence ($w$) | 23% | 17% | 60% | 20% | 15% | 30% |
| Assortativeness mixing with main partners ($\varepsilon_M$) | 0.72 | 0.31 | 0.87 | 0.75 | 0.14 | 0.88 |
| Assortativeness mixing with casual partners ($\varepsilon_C$) | 0.85 | 0.66 | 0.89 | 0.75 | 0.20 | 0.89 |
| Vaccination rate before 25 July 2022 | 0.04% | 0.03% | 0.05% | 0.04% | 0.03% | 0.05% |
| Efficacy old vaccine ($v_1 = \sigma_1$) | 57% | 50% | 65% | 56% | 51% | 65% |
| Efficacy new vaccine ($v_2 = \sigma_2$) | 85% | 80% | 90% | 82% | 80% | 90% |
| Hospitalization rate, $\zeta$ | 2% | 1% | 2% | 2% | 1% | 2% |
| Days infectious not refraining from sex, before $T_2$ | 6.20 | 2.73 | 7.87 | 5.82 | 4.62 | 7.98 |
| Days infectious not refraining from sex, after $T_2$ | 2.80 | 2.02 | 6.60 | 2.24 | 2.02 | 2.80 |
| Date of adaptations in July, $T_2$ | 8-7-2022 | 5-7-2022 | 14-7-2022 | 6-7-2022 | 5-7-2022 | 9-7-2022 |
| % reduction casual partners, low activity groups, after $T_2$ | 13% | 0% | 29% | 11% | 0% | 28% |
| % reduction casual partners, fairly-high activity group, after $T_2$ | 17% | 1% | 29% | 14% | 1% | 29% |
| % reduction casual partners, very-high activity group, after $T_2$ | 22% | 2% | 29% | 22% | 2% | 29% |

**Table S3.**
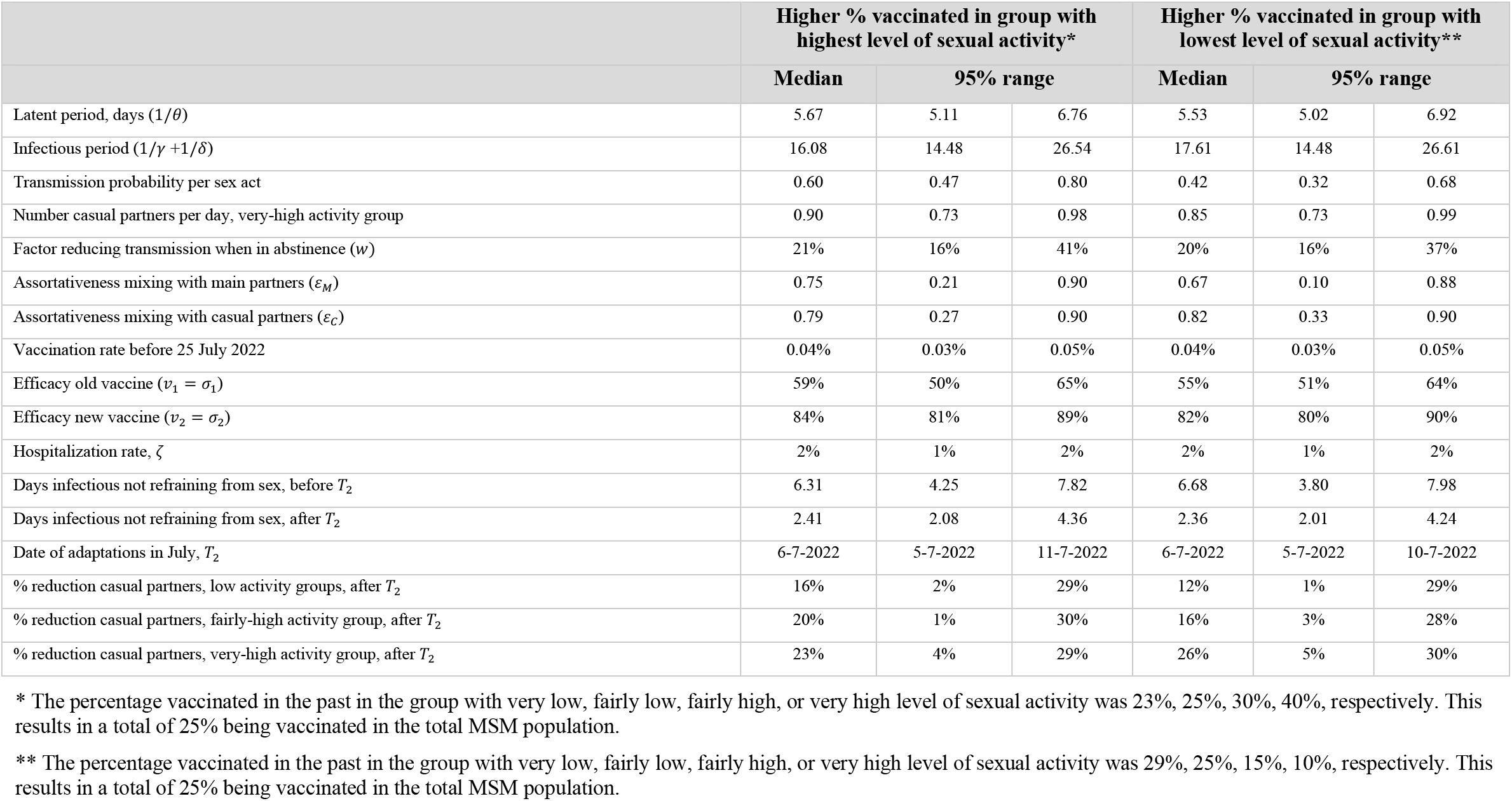
Sensitivity analysis for the percentage of MSM vaccinated in the past via the old smallpox vaccination programme. Posterior distributions of uncertain model parameters obtained from the fitting process for the scenario with adaptations in behaviour of MSM only in July 2022. Results shown are the medians and 95% credible intervals, in a population of 250,000 MSM.

|  | Higher % vaccinated in group with highest level of sexual activity* |  |  | Higher % vaccinated in group with lowest level of sexual activity** |  |  |
| --- | --- | --- | --- | --- | --- | --- |
|  | Median | 95% range |  | Median | 95% range |  |
| Latent period, days ($1/\theta$) | 5.67 | 5.11 | 6.76 | 5.53 | 5.02 | 6.92 |
| Infectious period ($1/\gamma + 1/\delta$) | 16.08 | 14.48 | 26.54 | 17.61 | 14.48 | 26.61 |
| Transmission probability per sex act | 0.60 | 0.47 | 0.80 | 0.42 | 0.32 | 0.68 |
| Number casual partners per day, very-high activity group | 0.90 | 0.73 | 0.98 | 0.85 | 0.73 | 0.99 |
| Factor reducing transmission when in abstinence ($w$) | 21% | 16% | 41% | 20% | 16% | 37% |
| Assortativeness mixing with main partners ($\varepsilon_M$) | 0.75 | 0.21 | 0.90 | 0.67 | 0.10 | 0.88 |
| Assortativeness mixing with casual partners ($\varepsilon_C$) | 0.79 | 0.27 | 0.90 | 0.82 | 0.33 | 0.90 |
| Vaccination rate before 25 July 2022 | 0.04% | 0.03% | 0.05% | 0.04% | 0.03% | 0.05% |
| Efficacy old vaccine ($v_1 = \sigma_1$) | 59% | 50% | 65% | 55% | 51% | 64% |
| Efficacy new vaccine ($v_2 = \sigma_2$) | 84% | 81% | 89% | 82% | 80% | 90% |
| Hospitalization rate, $\zeta$ | 2% | 1% | 2% | 2% | 1% | 2% |
| Days infectious not refraining from sex, before $T_2$ | 6.31 | 4.25 | 7.82 | 6.68 | 3.80 | 7.98 |
| Days infectious not refraining from sex, after $T_2$ | 2.41 | 2.08 | 4.36 | 2.36 | 2.01 | 4.24 |
| Date of adaptations in July, $T_2$ | 6-7-2022 | 5-7-2022 | 11-7-2022 | 6-7-2022 | 5-7-2022 | 10-7-2022 |
| % reduction casual partners, low activity groups, after $T_2$ | 16% | 2% | 29% | 12% | 1% | 29% |
| % reduction casual partners, fairly-high activity group, after $T_2$ | 20% | 1% | 30% | 16% | 3% | 28% |
| % reduction casual partners, very-high activity group, after $T_2$ | 23% | 4% | 29% | 26% | 5% | 30% |

\* The percentage vaccinated in the past in the group with very low, fairly low, fairly high, or very high level of sexual activity was 23%, 25%, 30%, 40%, respectively. This results in a total of 25% being vaccinated in the total MSM population.
\*\* The percentage vaccinated in the past in the group with very low, fairly low, fairly high, or very high level of sexual activity was 29%, 25%, 15%, 10%, respectively. This results in a total of 25% being vaccinated in the total MSM population.

